# The Mediating Role of Sleep in the Association Between Environmental Noise and Mental Health

**DOI:** 10.1101/2024.07.02.24309814

**Authors:** Kaya Grocott, Adelle Mansour, Rebecca Bentley, Kate E. Mason

## Abstract

Exposure to environmental noise in residential areas has been associated with adverse mental health outcomes; however, the mechanisms of this relationship remain underexplored. This study investigates the contribution of reduced sleep quality to the negative association between perceived neighbourhood environmental noise exposure and poor mental health. We used the Household Income and Labour Dynamics in Australia (HILDA) survey and applied causal mediation methods to examine the role of sleep in the association between self-reported exposure to road traffic noise and plane, train and industry (PTI) noise and mental wellbeing at three time points between 2012 and 2021. Road traffic noise was associated with poorer mental health in 2012-13 and 2016-17, while no evidence of an association was observed in 2020-2021 (the period of COVID-related lockdowns in Australia). For the years where a significant association was observed, mediation analyses suggest that reduced sleep quality accounts for 21% (in 2012-13; 95% CI: 7–35%) and 33% (in 2016-17; 95% CI: 26–64%) of the total effect of perceived traffic noise on mental health. Perceived PTI noise was associated with poorer mental health in 2016-2017 and 2020-2021, with mediation through sleep observed in 2016-2017 (proportion mediated 20% (95% CI:3–38%)). Mediation by sleep quality was stronger among people reporting exposure to multiple noise sources than among people reporting exposure to a single noise source. As much as a third of the association between road traffic noise and poor mental wellbeing may be due to poorer sleep quality following exposure to unwanted noise.

## INTRODUCTION

Environmental noise, defined as unwanted or disruptive outdoor sound caused by human activities, can adversely affect the health and wellbeing of individuals and is becoming increasingly pervasive due to growing urbanisation[1,2]. Previous studies estimate that in western Europe environmental noise accounts for the loss of 1.0-1.6 million disability-adjusted life years (DALYs) annually, primarily due to its effects on health and wellbeing, including childhood cognitive impairment, sleep disturbance, tinnitus and noise-annoyance[3]. Environmental noise stems from several sources, ranging from transport (road, rail, air) and industry to recreation and domestic activities[3]. With an expanding proportion of the global population living in urban areas, understanding the health impact of environmental noise exposure in residential areas is important[4].

Some research suggests chronic exposure to environmental noise is also associated with increased stress, anxiety and depressive symptoms, although the quality of the currently available evidence is generally low and results are heterogeneous [5,6]. Most studies rely on cross-sectional data, and limited attention has been given to understanding the mediating factors that would explain these associations. The most thoroughly studied mediator of the relationship between noise and mental health is noise annoyance[7–11]. Among lesser understood candidate mediators is sleep, which plays a crucial role in maintaining both physical and mental wellbeing[12]. Improved sleep quality is known to contribute to good mental health[13], yet night-time environmental noise can adversely impact sleep[14]. It can affect individual’s sleep quality, causing them to awaken or preventing them from falling asleep entirely[14]. Therefore, it is highly plausible that adverse mental health effects associated with exposure to noise in the neighbourhood environments surrounding people’s homes are mediated by reduced sleep quality. The small number of studies investigating sleep as a mediator of the relationship between noise pollution and mental health are narrowly focussed, concentrating either on university student populations[8–10] or a specific noise source (wind turbines[7]), and therefore may not be generalisable to the wider population.

This study uses nationally representative panel data from a large sample of Australian households to explore whether sleep mediates the association between environmental noise exposure and mental health among the community-dwelling population aged 15 and above. Examining this issue in the Australian context is important since the urban population accounts for approximately 70% of the total population[15], and the nation trails behind other regions with regard to understanding the impact that exposure to environmental noise has on the community[16]. The panel structure of the data used allows us to establish appropriate temporal ordering of the exposure, potential mediators and the outcome.

We focus on perceived noise from road traffic and plane, train, and industry (PTI) noise: sources that make up much of the environmental noise in residential areas[17]. Prior studies have shown that perceived noise is associated with psychological stress and other mental health outcomes[18,19]. While objective noise measurement is critical for evaluating the relationship between environmental noise exposure and health[20], subjective experiences of noise are also important. Subjective experiences of sound differ among individuals – the same sound may be deemed as noise by some and not by others[21]. In the context of mental health effects in particular, therefore, considering measures of perceived noise can contribute to a more nuanced understanding of the impacts of noise in the residential environment.

By exploring the role of sleep as a potential mediator, we aim to improve current understanding of the pathways through which transport and industry noise in the residential environment impacts mental wellbeing, thereby contributing to the development of interventions and policies to reduce environmental noise exposure and, ultimately, promote healthier communities.

## METHODS

### Study Population

This study used the Household Income and Labour Dynamics in Australia (HILDA) survey, a longitudinal study running since 2001. HILDA follows Australians over the course of their lives and collects data on households, family and relationships, financial wellbeing, participation in work and education and health. It has previously been used to explore the relationship between environmental noise and mental wellbeing[19]. The sampling unit is the household, whose members are followed up annually. Using a multistage clustered, stratified design HILDA commenced with a nationally representative sample of 7,682 Australian households and 13,969 people and has gradually expanded to include new members of original households, new households formed by original participants, and a top-up sample in wave 11. The 2021 sample (wave 21) consisted of 9,358 households with survey responses from 16,549 people[22]. The HILDA dataset is deidentified and publicly available to approved researchers through the Australian Government Department of Social Services (https://dataverse.ada.edu.au/dataverse/DSSLongitudinalStudies). As such this study was exempt from institutional review board approval.

### Measures

#### Mental health outcome

Mental health was measured annually in HILDA using the mental health component summary score of the validated Short Form 36 (SF-36) survey[23]. The SF-36 survey is a self-rated health survey providing a comprehensive assessment that distinguishes between mental and physical health[23]. The mental health summary score ranges from 0-100 with higher scores indicative of better mental health.

#### Noise exposure

The HILDA survey asks participants biennially about the frequency of noise in their neighbourhood: specifically, plane, train and industry (PTI) noise and road traffic noise. Responses were recorded as 1 (never happens), 2 (very rare), 3 (not common), 4 (fairly common) and 5 (very common). For analysis, we dichotomised noise exposure such that 1 indicated fairly/very common noise exposure and 0 represented less frequent noise. We analysed road traffic and PTI noise both separately and in combination. For the latter, we derived an additional exposure variable to indicate whether each participant was exposed fairly/very commonly to 0 (no noise), 1 (road traffic noise only), 2 (PTI noise only) or 3 (both).

#### Sleep quality

Sleep quality has been measured in HILDA every four years since 2013. Participants rate their sleep over the last 30 days on a scale from 1 (very good) to 4 (very bad). This variable was dichotomised for analysis, with 1 indicating fairly/very bad sleep quality and 0 indicating fairly/very good sleep.

#### Covariates

We controlled for a range of demographic, socioeconomic, health and housing covariates; specifically, sex, baseline mental health, labour force status, shiftwork status, household income, education, alcohol consumption, several major life events, area disadvantage and whether residing in a major city. The relative timing of the measurement used for each covariate was determined by whether it was considered a potential mediator of the exposure-outcome, exposure-mediator, and/or mediator-outcome association. See Appendix 1 for covariate details.

### Statistical Analysis

Data were analysed in Stata/SE 18.0 for three 1-year follow up periods: 2012-13, 2016-17 and 2020-21. Analyses were repeated for each period, with noise and baseline covariates measured in the first year and sleep and mental health measured the following year. Binary life event indicators were based on events reported in the second year, to capture events that occurred after the measurement of noise and in the year leading up to the measurement of sleep and mental health, to ensure appropriate temporal sequencing as these were considered potential confounders only of the mediator-outcome association. The analytic samples were restricted to individuals who did not experience a residential move during follow up, to minimise the chance of a substantial change in noise exposure before sleep and mental health were measured. Complete case analyses were performed.

Figure 1 depicts the relationships we investigated in our analyses. For each exposure (road traffic, PTI and combined noise), we first estimated its association with mental health (using multiple linear regression) and, separately, sleep quality (using multiple logistic regression), after adjusting for potential confounders. We also used estimated associations between sleep quality and mental health. We then used the ‘mediate’ package in Stata 18 to estimate total, direct and natural indirect effects of noise exposure on mental health, and proportion mediated, with sleep quality as the potential mediator. This approach is based on the potential-outcomes framework that considers observed outcomes in relation to a counterfactual scenario of no exposure. Causal mediation is more robust than traditional mediation methods such as the difference method because it relies on fewer assumptions; in particular causal mediation allows for potential exposure-mediator interactions and non-linear relationships rather than assuming these are not present [24,25]. As the presence of an exposure-mediator interaction was unknown from prior research, we also tested for these to assess whether our decision to use a model that relaxes this assumption was important for this relationship.

**Figure 1.**
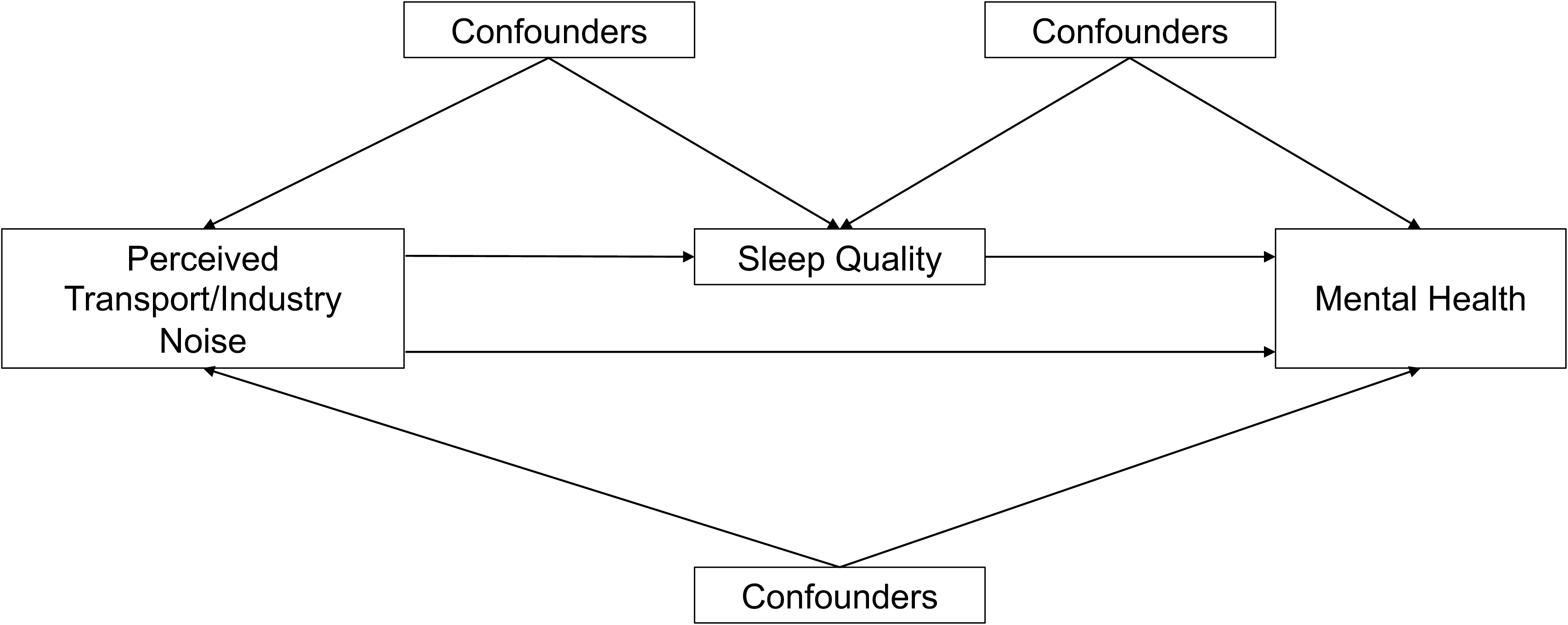
DAG

### Sensitivity analyses

To examine the sensitivity of our results to the way noise exposure was defined, we repeated the analysis redefining the exposed category to only those individuals exposed to noise “very commonly”. This also allowed us to examine whether sleep quality is a stronger mediator in individuals exposed to more frequent noise.

## RESULTS

### Summary statistics

Table 1 describes the sample of respondents for each follow-up period (2012-2013, 2016-2017 and 2020-2021) with sample sizes of 10,605, 11,536 and 10,991 respectively. The proportion of participants reporting good sleep quality decreased slightly from 75.7% in 2013 to 71.7% in 2021. Noise exposure for both road traffic noise and PTI noise remained relatively stable across the three time periods, with an average of 26.8% of individuals reporting fairly/very common exposure to road traffic noise and 22% reporting fairly/very common exposure to PTI noise.

**Table 1:**
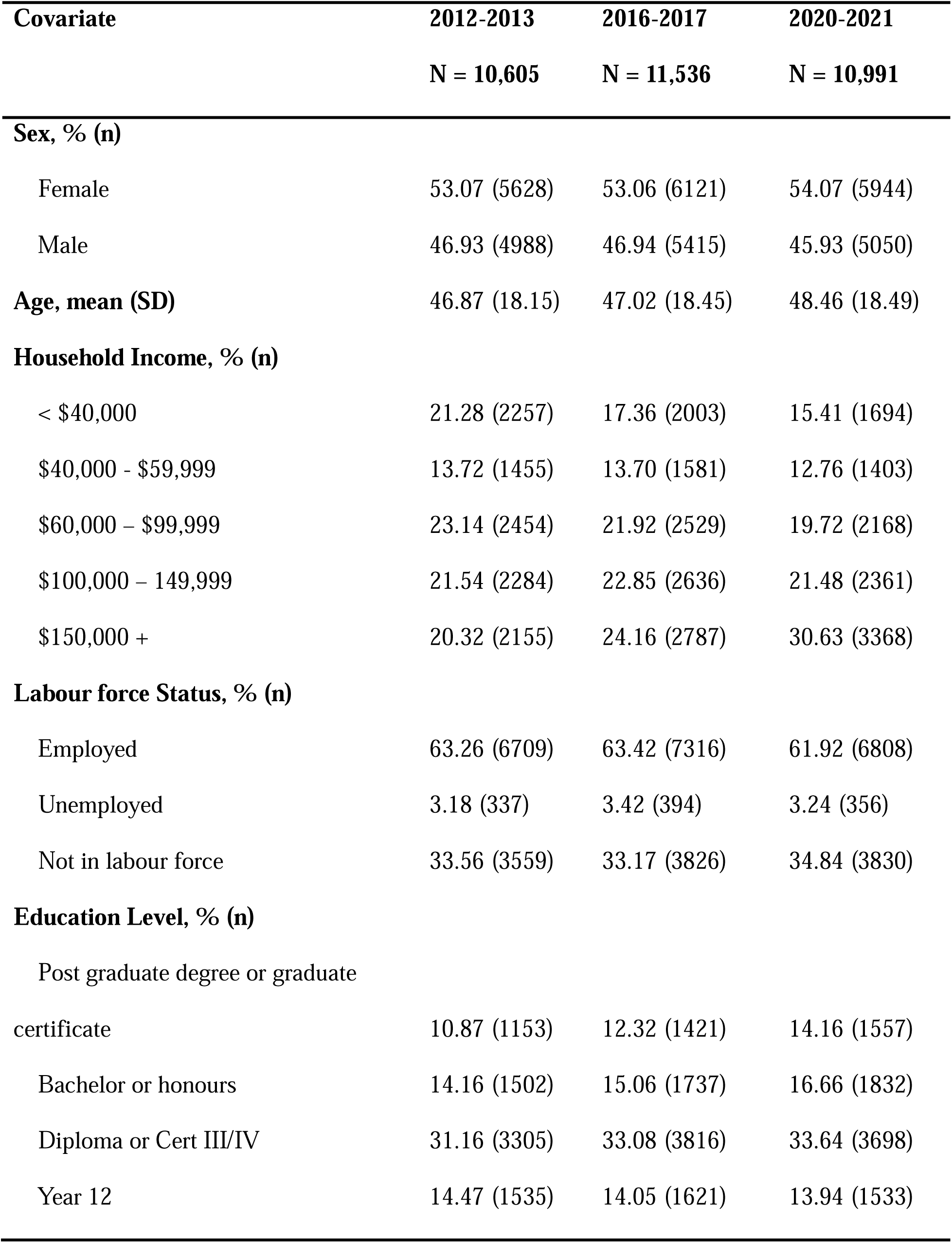
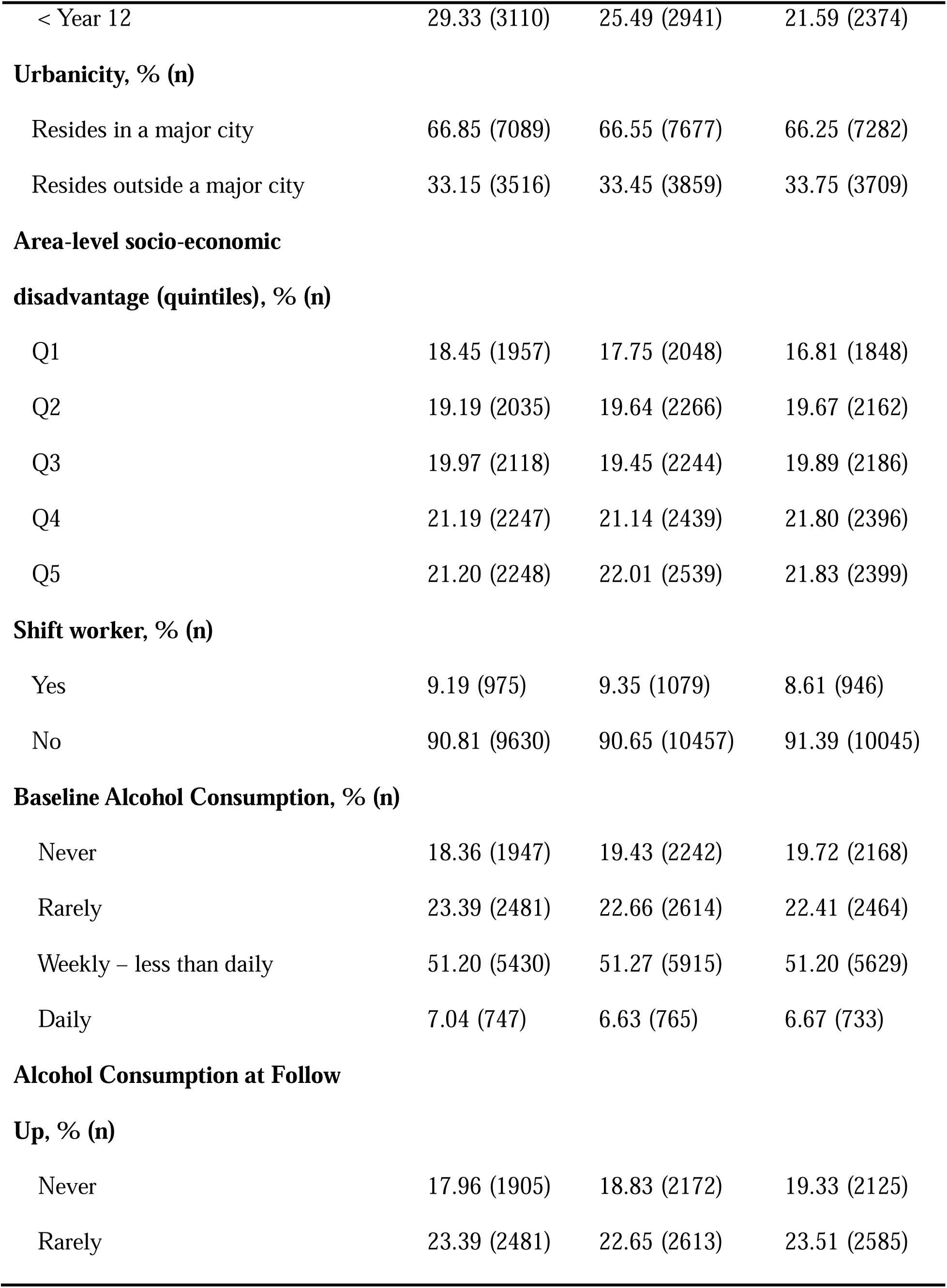
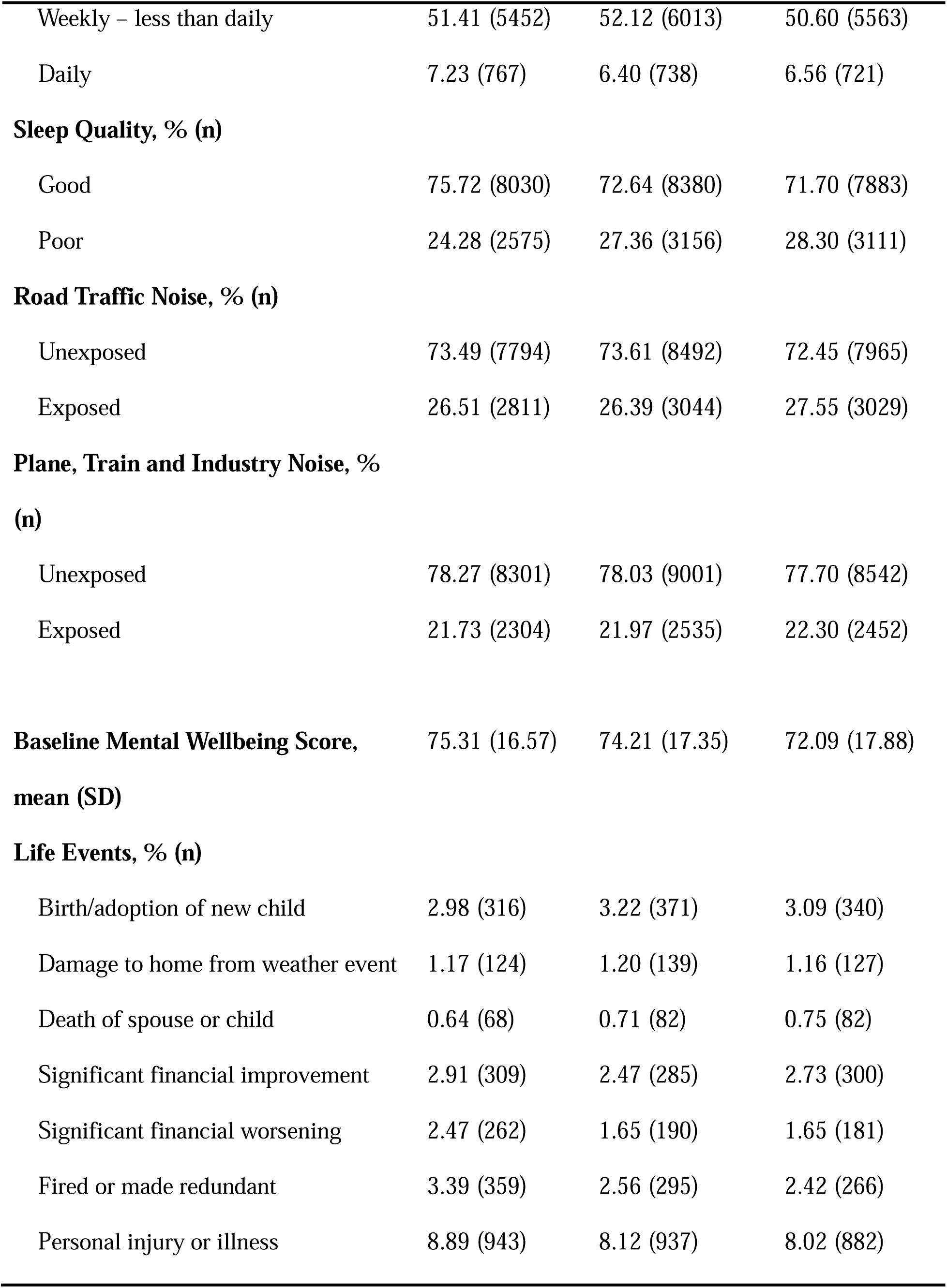
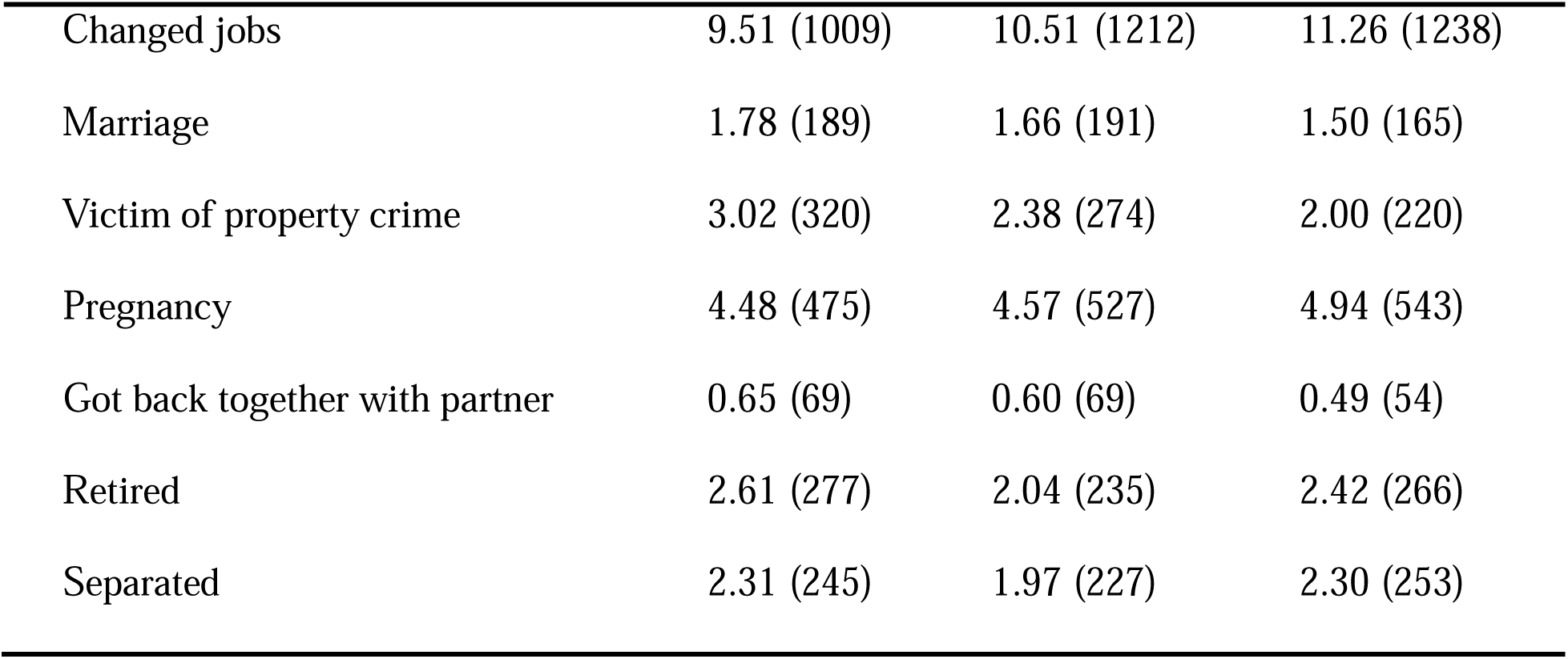
Characteristics of Study Sample.

### Associations between noise, sleep and mental health

Frequent exposure to both road traffic and PTI noise were consistently associated with poorer mental health, although the magnitude of the association varied between years (Appendix Table 2). Road traffic noise was more strongly associated with mental health than was PTI noise in 2012-2013 (road traffic: β = −1.17; 95% CI: −1.72, −0.63; PTI noise β = −0.22; 95% CI: −0.80, 0.35), while the effect of PTI noise on mental health appeared greater in 2016-2017 (road traffic: β = −0.70; 95% CI: −1.23, −0.16; PTI noise β = −0.82; 95% CI: −1.39, −0.26) and 2020-2021 (road traffic: β = −0.32; 95% CI: −0.86, 0.22; PTI noise β = −0.48; 95% CI: −1.06, 0.09). Road traffic noise was associated with increased odds of poor sleep in 2012-2013 (OR = 1.21; 95% CI: 1.10, 1.35) and 2016-2017 (OR = 1.18; 95% CI: 1.07, 1.30) (Appendix Table 3). PTI noise was associated with increased odds of poor sleep quality in 2016-2017 only (OR = 1.19; 95% CI: 1.07, 1.31). There was no evidence to support an association between any source of noise exposure and sleep quality in 2020-21. A strong negative association between sleep quality and mental health was observed for all years (2012-13: β = −6.11; 95% CI: −6.67, −5.55; 2016-17: β = −6.27; 95% CI: −6.79, −5.74; 2020-21: β = −5.80; −6.34, −5.26) (Appendix Table 4).

### Mediation analysis

Causal mediation analysis showed a strong negative indirect effect of road traffic noise on mental health through sleep quality (Table 2) in both 2012-13 (natural indirect effect (NIE) = −0.22; 95% CI: −0.34, −0.09) and 2016-17 (NIE = −0.21; 95% CI: −0.34, −0.-0.08). The proportions mediated were estimated to be 21% (7%, 35%) and 33% (26%, 64%) respectively. In 2020-2021 there was little to no evidence of an association between road traffic noise exposure and mental health and therefore sleep could not act as a mediator, though the estimated effect sizes for the total and indirect effect were consistently negative.

**Table 2:** Mental Health Impact of Single source Noise Mediated by Sleep Quality.

|  | **Total Effect** | **P-**<br>**value** | **Natural Indirect**<br>**Effect** | **P-value** | **Natural Direct**<br>**Effect** | **P-**<br>**value** | **Proportion**<br>**Mediated** | **P-value** |
| --- | --- | --- | --- | --- | --- | --- | --- | --- |
| **Road Traffic Noise** |  |  |  |  |  |  |  |  |
| 2012-13 | **-1.06 (-1.6, -0.50)** | **<0.001** | **-0.22 (-0.34, -0.09)** | **0.001** | **-0.84 (-1.38, -0.30)** | **0.002** | **0.21 (0.07, 0.35)** | **0.004** |
| 2016-17 | **-0.63 (-1.18, -0.09)** | **0.023** | **-0.21 (-0.34, -0.08)** | **0.001** | -0.42 (-0.95, 0.11) | 0.118 | **0.33 (0.26, 0.64)** | **0.034** |
| 2020-21 | -0.19 (-0.75, 0.36) | 0.500 | -0.06, (-0.17, 0.05) | 0.311 | -0.13 (-0.67, 0.41) | 0.631 | 0.31 (-0.65, 1.26) | 0.531 |
| **Plane Train and Industry Noise** |  |  |  |  |  |  |  |  |
| 2012-13 | -0.10 (-0.67, 0.48) | 0.740 | -0.08 (-0.21, 0.05) | 0.206 | -0.01 (-0.57, 0.54) | 0.959 | 0.85 (-4.01, 5.73) | 0.734 |
| 2016-17 | **-0.81 (-1.38, -0.25)** | **0.005** | **-0.17 (-0.27, -0.06)** | **0.002** | **-0.65 (-1.20, -0.09)** | **0.022** | **0.20 (0.03, 0.38)** | **0.019** |
| 2020-21 | -0.45 (-1.04, 0.13) | 0.130 | -0.03 (-0.15, 0.09) | 0.596 | -0.42 (-1.00, 0.15) | 0.150 | 0.07 (-0.19, 0.32) | 0.592 |

Adjusted for age, sex, baseline mental health, household income, labour force status, education level, area socioeconomic disadvantage, shift work, residence in a major city, alcohol consumption at baseline and follow up and several life events. Boldface indicates statistical significance (p<0.05).

The indirect effect of PTI noise on mental health was less consistent across the years, however all years had a negative indirect effect estimate (Table 2). The strongest indirect effect was observed in 2016-17 (NIE = −0.17; 95% CI: −0.27, −0.06), accounting for 20% (3%, 38%) of the total effect. No association between PTI noise and mental health was observed in 2012-2013 or 2020-2021.

When combined exposure to both PTI noise and road traffic noise was compared to exposure to only one source of noise, the indirect effect of noise on mental health via sleep quality was larger in those exposed to multiple noise sources than either source individually, in both 2012-13 and 2016-17 (Table 3). Consistent with results when noise sources were analyses separately, no total or mediated effects were observed in 2020-21 for combined or lone source noise exposure.

**Table 3.**
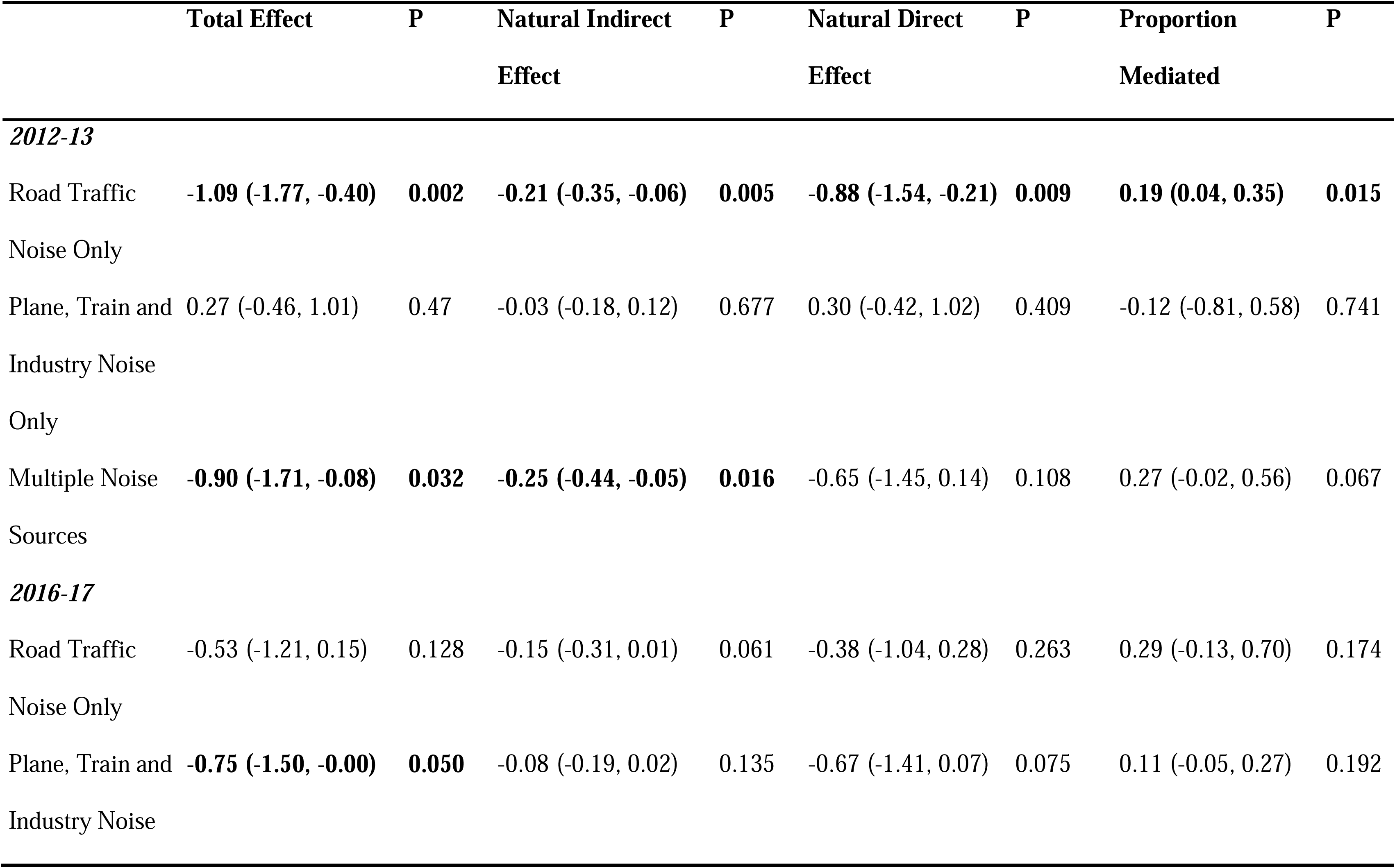
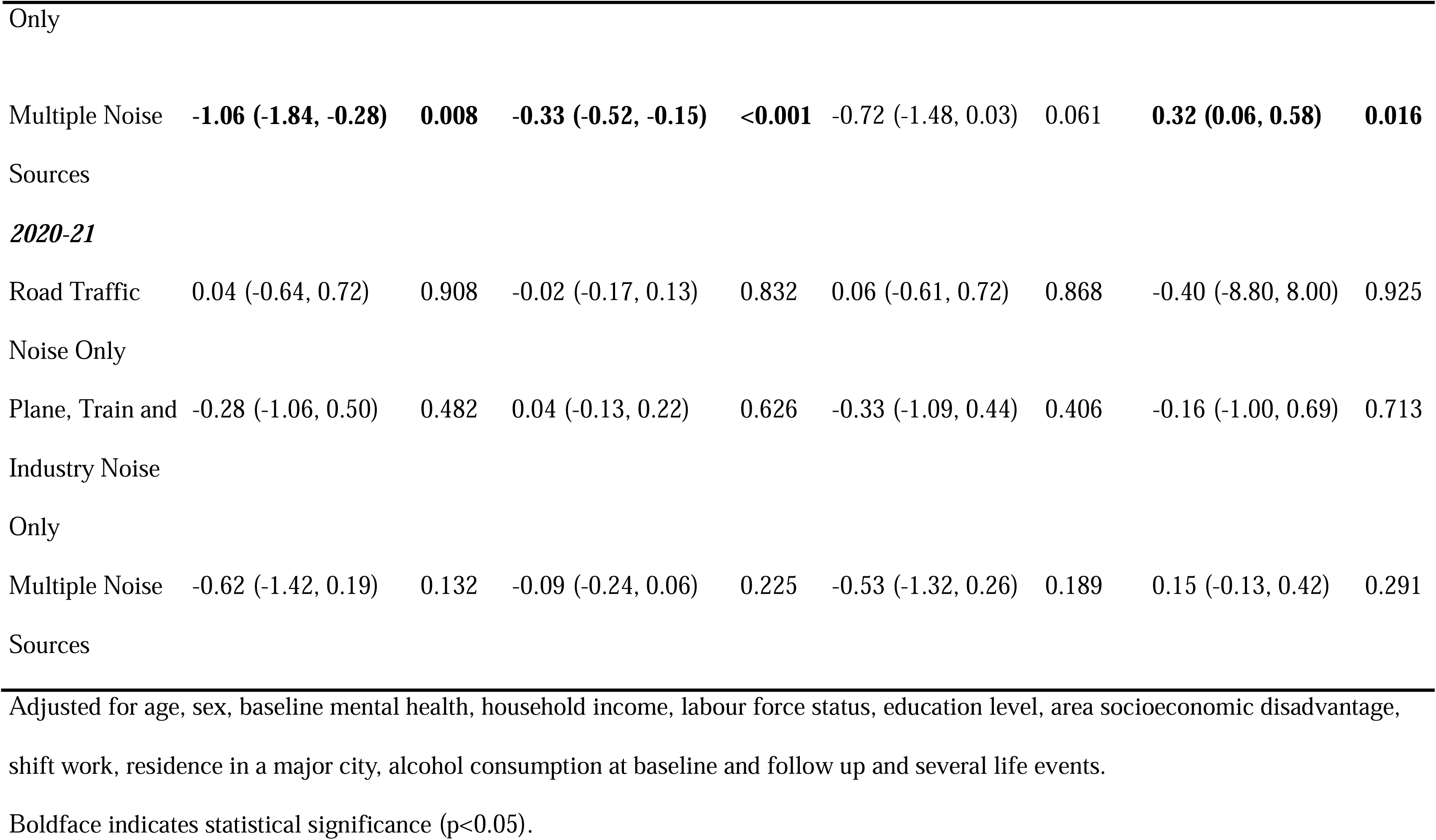
Mental Health Impact of Multi- and Single-Source Noise Exposure Mediated by Sleep.

There was only minimal evidence of exposure-mediator interaction (Appendix 4), any effect of which will have been accounted for by using a causal mediation modelling approach.

### Sensitivity Analyses

In the sensitivity analysis, when the exposed category was limited to those exposed very commonly to noise, results for both road traffic noise and PTI noise were broadly consistent with the primary analysis (Appendix Table 5).

## DISCUSSION

Our study provides evidence that perceived road traffic noise exposure in the neighbourhood adversely impacts mental health, with sleep quality appearing to mediate the association in both 2012-13 and 2016-17. In contrast, in 2020-21 evidence for any association between road traffic noise exposure and mental health was limited, despite perceived road traffic noise levels remaining stable. With much of Australia subject to COVID-19 lockdowns during 2020-21, it may be that our findings for that period were influenced by a combination of increased time at home, changes in objective levels of noise from various sources, working/schooling from home, and competing influences on mental health at that time.

The association between mental health and self-reported noise from planes, trains and industry (as opposed to road traffic) was inconsistent across years and across our various sensitivity analyses, leading us to conclude that any mental health effect of PTI noise is likely to be weaker than the effect of road traffic noise. However, results of our mediation analysis when PTI was associated with mental health (in 2016-17 only) show evidence of an indirect pathway through sleep, suggesting that if PTI noise does influence mental health, the relationship may be mediated by sleep. When considering perceived exposure to multiple noise sources compared to exposure to a single source of noise, multiple exposures also had larger sleep-mediated effects on mental health.

Our findings are consistent with numerous previous studies reporting a negative association between both perceived and actual traffic noise exposure in the neighbourhood and mental health[19,26–29] and align with prior research concluding that mental health was less impacted by PTI noise than road traffic noise[27]. Studies have shown that at the same levels of noise railway noise is commonly perceived as less annoying than noise from road or air traffic however frequency of trains, building and vibration factors may influence this relationship [14,30]. One previous study showed that sleep is only a mediator when an individual also experiences some level of noise annoyance[26]; it is plausible that this lower level of annoyance may be sufficient to reduce the mental health impact of PTI noise[8]. Our results are also consistent with the findings from research using objectively measured noise in showing cumulative effects of multiple noise sources on mental health[27].

We are aware of four studies that have previously investigated the mediating role of sleep with respect to noise and mental health[7–10]. However, these have been limited in their scope and varied in results. Three of these studies were conducted in university students and, like the current study, found sleep to be a mediator of the association between noise and mental health, though one found this was only the case in individuals reporting noise annoyance. The remaining study[7] conversely found no mediating effect of sleep, however it was limited to noise exposure from wind turbines. Building on this limited existing evidence base, our study’s strengths lie in its longitudinal study design and large sample size allowing for a comprehensive exploration of the mediating role of sleep in the noise-mental health relationship in the general adult population. Repeating our analysis over three separate time periods introduced replication, enhancing the credibility of the findings.

The study nonetheless has some limitations. First, in using self-reported measures of noise exposure we are limited to perceptions of noise rather than objectively measured exposure, and risk introducing bias due to measurement error. However, a previous study using the same dataset employed an area-level aggregate measure of perceived environmental noise to reduce bias from individual-level measurement error and found that associations with mental health were of a similar magnitude to when using the individual measure [19]. Second, the study’s exclusive focus on transport and industry noise within the neighbourhood environment omitted noise from neighbours and elsewhere within the neighbourhood as well as possible occupational and commuting noise exposure. Third, we were also not able to consider the amount of time spent at home, and thereby assume all individuals are equally exposed to the noise in their home environment, when in truth differential exposure is likely and may have led us to underestimate importantly larger effects in some groups of people. Relatedly, the source data also did not allow us to consider the timing of noise exposures during the day/night, which may be vital in understanding the impact of noise on sleep. Fourth, we relied on a single item self-reported measure of sleep, as more comprehensive sleep measurement was not available. Future research is needed using more robust measurement of sleep (e.g. validated sleep questionnaires and actigraphy) as part of this relationship. Finally, our models did not include noise sensitivity. As noise sensitivity could lead to increased reporting of environmental noise exposure and can be increased by poor mental health, this omission introduces the possibility of reverse causation[12,31].

Overall, our findings suggest that strategies aimed at improving sleep quality, especially in areas impacted by high levels of environmental noise from multiple sources, have the potential to improve population mental health. This could extend to the design of homes, and particularly bedrooms, in aiming to protect against sleep disturbance caused by noise. Upstream interventions aimed at reducing environmental noise in residential areas should consider the differential impact of noise from various sources. Road traffic noise appears more harmful that PTI noise though areas with high levels of both types of noise may place residents at the highest risk.

Future research should explore the role of sleep quality as a mediator in the context of different noise sources and using validated measures of perceived and objectively measured noise and sleep. Additionally, further consideration should be given to potential effect modification by co-occurring exposures such as air and light pollution, and housing conditions and design features (e.g. insulation, layout), which are likely to moderate or compound effects of noise on sleep and mental health.

## Conclusions

This study provides evidence to support the role of sleep quality in explaining the association between noise exposure in the neighbourhood and mental health. We found that sleep quality accounts for one-fifth to one-third of the estimated total effect of noise on mental health among Australian adults. At present this is the only study to examine the mediating effects of sleep on the mental health impact of environmental noise using national, longitudinal data. Further longitudinal studies that make use of validated measures are warranted. Interventions that target the layouts of neighbourhoods, transport management, and the design and construction of homes to reduce the impact of noise exposures on sleep quality have the potential to improve population-level health and wellbeing.

## Data Availability

Data from the HILDA Survey is available to researchers living in Australia or overseas through the Australian Government Department of Social Services Longitudinal Studies Dataverse.

https://dataverse.ada.edu.au/dataverse/DSSLongitudinalStudies

