## Supplementary material for "The Mediating Role of Sleep in the Association Between Environmental Noise and Mental Health"

#### 1. Covariates

Covariates controlled for in this analysis were, age in years, sex, baseline mental health measured by the SF-36 survey, labour force status (employed/unemployed/ not in labour force), household income (<$40,000/ $40,000 - $59,999/$60,000-$99,999/$100,000-$149,999/$150,000), area socioeconomic disadvantage (quintiles of the Australian Bureau of Statistics' Socio-Economic Index for Areas (SEIFA) 2011 Index of relative socio-economic advantage/disadvantage), shift worker (yes/no, where shift work includes regular evening or night shifts, rotating day/night shifts, split shifts, or being on call), residence in a major city (yes/no), education level (Post-graduate studies/ Undergraduate studies/Diploma or Cert III-IV/ year 12/< year 12), alcohol consumption at baseline and follow-up (Never, rarely, weekly- less than daily, daily), and binary indicators for the birth/adoption of a child, weather related damage to home, death of spouse or child, major financial improvement or worsening, job loss, personal illness or injury, job changes, being the victim of a property crime, pregnancy, changes in relationship status including marriage, separating and reconciling with a partner and retirement.

**Appendix Table 1 Variable Availability Across Waves, 2013-2021**

| | 2012 | 2013 | 2014 | 2015 | 2016 | 2017 | 2018 | 2019 | 2020 | 2021 |
|---|---|---|---|---|---|---|---|---|---|---|
| Noise | x | | x | | x | | x | | x | |
| Sleep | | x | | | | x | | | | x |
| SF36 MH | x | x | x | x | x | x | x | x | x | x |
| Covariates | x | x | x | x | x | x | x | x | x | x |

#### 2. Associations Between Noise, Sleep and Mental Health

**Appendix Table 2 Associations Between Self-Reported Noise and Mental Health**

| | Road Traffic Noise | P-value | Plane, Train and Industry Noise | P-value |
|---|---|---|---|---|
| 2012-2013 | **-1.17 (-1.72, -0.63)** | **< 0.001** | -0.22 (-0.80, 0.35) | 0.448 |
| 2016-2017 | **-0.70 (-1.23, -0.16)** | **0.011** | **-0.82 (-1.39, -0.26)** | **0.004** |
| 2020-2021 | -0.32 (-0.86, 0.22) | 0.249 | -0.48 (-1.06, 0.09) | 0.101 |

Note: Linear regression models adjusted for age, sex, baseline mental health, household income, labour force status, education level, alcohol consumption at baseline, area socioeconomic disadvantage, shift work, residence in a major city, and several life events. Boldface indicates statistical significance ($p<0.05$).

**Appendix Table 3 Relative Odds of Sleep Quality with Noise Exposure**

| | Road Traffic Noise (OR, 95% CI) | P-value | Plane, Train and Industry Noise (OR, 95% CI) | P-value |
|---|---|---|---|---|
| 2012-2013 | **1.21 (1.10, 1.35)** | **< 0.001** | 1.08 (0.96, 1.20) | 0.197 |
| 2016-2017 | **1.18 (1.07, 1.30)** | **0.001** | **1.19 (1.07, 1.31)** | **0.001** |
| 2020-2021 | 1.05 (0.95, 1.16) | 0.307 | 1.03 (0.93, 1.14) | 0.595 |

Note: Logistic regression models adjusted for age, sex, baseline mental health, household income, labour force status, education level, alcohol consumption at baseline, area socioeconomic disadvantage, shift work, residence in a major city, and several life events. Boldface indicates statistical significance ($p<0.05$).

**Appendix Table 4 Associations Between Self-Reported Sleep Quality and Mental Health**

| | Sleep Quality | P-value |
|---|---|---|
| 2012-2013 | **-6.11 (-6.67, -5.55)** | **< 0.001** |
| 2016-2017 | **-6.27 (-6.79, -5.74)** | **< 0.001** |
| 2020-2021 | **-5.80 (-6.34, -5.26)** | **< 0.001** |

Note: Linear regression models adjusted for age, sex, baseline mental health, household income, labour force status, education level, alcohol consumption at follow up, area socioeconomic disadvantage, shift work, residence in a major city, and several life events. Boldface indicates statistical significance ($p<0.05$).

#### 3. Sensitivity Analyses

A sensitivity analysis was conducted by changing the definition of exposed for both traffic and PTI noise, such that only individuals who reported “very common” exposure to noise were classified as exposed to noise (as opposed to “very common” and “fairly common” in the primary analysis). For road traffic noise, the total and indirect effect estimates, and proportions mediated, were similar to those from the primary analysis across all years, but point estimates were less precise (Appendix Table 5). For PTI noise, evidence for total and mediated effects were seen only for 2016-17 in the primary analysis. With the narrower definition of exposed, the less precise estimates contributed to weaker evidence for that period. Consistent with primary analysis, no associations or mediation were observed for PTI noise in the other years.

**Appendix Table 5 Sensitivity Analysis: Exposure redefined as only "very common" exposure to noise**

| | **Total Effect** | **P-value** | **Natural Indirect Effect** | **P-value** | **Natural Direct Effect** | **P-value** | **Proportion Mediated** | **P-value** |
|---|---|---|---|---|---|---|---|---|
| **Road Traffic Noise** | | | | | | | | |
| **2012-13** | **-1.03 (-1.92, -0.14)** | **0.023** | **-0.23 (-0.43, -0.03)** | **0.021** | -0.80 (-1.66, 0.06) | 0.067 | 0.22 (-0.01, 0.46) | 0.061 |
| **2016-17** | 0.82 (-1.70, 0.06) | 0.069 | **-0.26 (-0.46, -0.06)** | **0.010** | -0.56 (-1.41, 0.30) | 0.204 | 0.32 (-0.05, 0.70) | 0.094 |
| **2020-21** | -0.11 (-1.06, 0.84) | 0.825 | -0.05 (-0.22, 0.12) | 0.573 | -0.06 (-0.99, 0.87) | 0.903 | 0.46 (-3.59, 4.51) | 0.824 |
| **Plane, Train and Industry Noise** | | | | | | | | |
| **2012-13** | -0.08 (-1.00, 0.84) | 0.872 | -0.20 (-0.42, 0.01) | 0.064 | 0.12 (-0.77, 1.02) | 0.872 | not estimated* | |
| **2016-17** | **-0.95 (-1.86, -0.03)** | **0.042** | -0.14 (-0.31, 0.03) | 0.099 | -0.81 (-1.70, -0.09) | 0.077 | 0.15 (-0.05, 0.35) | 0.151 |
| **2020-21** | -0.70 (-1.78, 0.35) | 0.190 | -0.21 (-0.44, 0.01) | 0.065 | -0.49 (-1.52, 0.53) | 0.347 | 0.30 (-0.19, 0.79) | 0.227 |

Note: Linear regression models adjusted for age, sex, baseline mental health, household income, labour force status, education level, alcohol consumption at baseline and follow up, area socioeconomic disadvantage, shift work, residence in a major city, and several life events.

Boldface indicates statistical significance ($p<0.05$).

* An interpretable proportion mediated could not be estimated for the PTI noise model in 2013-13 because signs among indirect, direct, and total effects were opposing.

### 4. Interactions

**Interactions**

Minimal evidence of interaction between perceived noise exposure and sleep was found (Appendix Table 6). Causal mediation analysis makes no assumptions about whether exposure-mediator interaction is present; in this instance, observing only limited evidence of interaction suggests our decision to use causal mediation analysis over traditional mediation methods may have been unimportant, with results from both approaches likely to be similar.

**Appendix Table 6 Exposure-mediator interactions in models of noise, sleep and mental health**

| | Noise-Sleep interaction term P-value | |
|---|---|---|
| | **Road Traffic Noise x Sleep Quality** | **Plane Train and Industry Noises** |
| **2012-2013** | 0.213 | 0.180 |
| **2016-2017** | 0.177 | **0.013** |
| **2020-2021** | 0.109 | 0.464 |

Note: Adjusted for age, sex, baseline mental health, household income, labour force status, education level and alcohol consumption at baseline and follow up, area socioeconomic disadvantage, shift work, residence in a major city, and several life events.
